# Identification of clinical, laboratory, and epidemiological characteristics to accelerate diagnosis of Crimean-Congo Hemorrhagic Fever in patients presenting to health centres in low-resource endemic settings: a systematic literature review

**DOI:** 10.1101/2025.09.15.25335745

**Authors:** Joseph Alberts, James Mason, Claire Mullender, Shamma Mumtaz

## Abstract

**Background:** Crimean-Congo Hemorrhagic Fever (CCHF) is a viral hemorrhagic fever caused by an orthonairovirus transmitted to humans by infected ticks. The incidence of CCHF is vastly underestimated, in part due to frequent misdiagnosis and poor surveillance infrastructure in endemic regions. The nonspecific presentation of CCHF makes diagnosis very challenging, especially in low-resource settings where diagnostic technologies, such as PCR, are scarce. The present study aimed to identify simple clinical, laboratory, and epidemiological characteristics that are predictive of CCHF diagnosis in the early stages of disease progression and have the potential to bridge this technological gap.

**Methodology:** A systematic search was conducted in Medline, Embase, Global Health, PubMed, and the Cochrane Library to identify eligible articles (PROSPERO ID: CRD42023434872). Studies comparing the characteristics of suspected CCHF-positive and CCHF-negative groups in countries where CCHF is endemic were included. Titles and abstracts were screened, with eligible full texts being reviewed by two reviewers. Included studies underwent data extraction of patient characteristics found to be present in significantly different levels in CCHF-positive patients. Odds ratios were calculated where possible.

The search identified 2,585 scientific studies, 14 of which were selected for this review. A total of 2,025 participants were included in the final analysis. Patient characteristics found to be most predictive of CCHF diagnosis were: transaminitis, headache, nausea/vomiting, CK elevation, fever, LDH elevation and recorded tick bites. In addition, leukopenia and thrombocytopenia were found to be especially predictive of CCHF diagnosis.

**Conclusions:** This review summarises characteristics of CCHF patients that may be of diagnostic utility. Similar findings in the literature support this study, although there are notable limitations concerning study design heterogeneity. These findings may be used to inform interim guidance until sensitive diagnostic technologies are made more available in remote/low-resource areas.

**Author Summary:** Crimean-Congo Hemorrhagic Fever (CCHF) is a serious infection caused by a virus that is spread through tick bites. It is often misdiagnosed as early symptoms are non-specific. Diagnostic tools are also limited in regions where the disease is common.

We carried out a systematic review of published studies to find out whether there are simple signs, symptoms, and routine blood test results that can help distinguish patients with this infection from other illnesses.

From over 2,000 studies identified, 14 were selected. The review found several key predictors of CCHF, including headache, nausea/vomiting, fever, elevated liver enzymes and recent tick bites. The strongest predictors of CCHF, however, were lower white blood cell and platelet counts in patients with CCHF.

Our findings bring together available evidence on practical indicators that could help frontline health workers recognise this neglected disease more quickly, especially in areas where advanced testing is not available.

## Introduction

Crimean-Congo Hemorrhagic Fever (CCHF) is a globally distributed, tick-borne viral zoonosis caused by the CCHF virus (of the Nairoviridae family) [1]. It is associated with significant mortality and morbidity, with a fatality rate as high as 40% recorded in outbreak settings [2].

CCHF is most prevalent in areas with frequent human-livestock interaction, as transmission occurs via contact with infected animal tissues or tick bites [3]. Human-to-human spread can occur through exposure to blood or bodily fluids, which poses a risk to healthcare workers [4]. Personal protective equipment (PPE) and stringent infection prevention and control (IPC) protocols in healthcare settings are crucial for limiting the spread of nosocomial infections [5].

There are over three billion people estimated to be at risk of CCHF infection globally, and between 10,000 and 15,000 infections are recorded across Africa, the Middle East, Asia, and increasingly, Europe [6]. The annual number of reported human cases is highly variable and likely underestimates the true incidence due to substantial underreporting and limited surveillance, especially in low- and middle-income countries.

Geographic distribution is expanding, linked with global warming and the dissemination of Hyalomma ticks. There is established endemicity in Russia, Turkey, the Balkans, and parts of Africa and Asia, and recent emergence of autochthonous cases in Greece, Spain, and Ukraine [7, 8].

Most human cases are asymptomatic, but the spectrum of disease severity is broad [9]. In the early (pre-hemorrhagic) phase, the clinical picture is often non-specific, with symptoms such as fever, myalgia, headache, nausea, vomiting, abdominal pain, and diarrhoea. The haemorrhagic phase, characterised by bleeding manifestations, typically develops after several days [10]. In low- and middle-income countries, these features appear very similar to other common causes of undifferentiated febrile illness. Misdiagnosis is frequent and has been reported in up to 68% of cases [11], leading to delayed recognition and increasing risk of transmission.

Definitive diagnosis requires Reverse Transcription-Polymerase Chain Reaction (RT-PCR) during, and/or Enzyme-Linked Immunosorbent Assay (ELISA). Viral RNA is only detectable in the first nine days (pre-haemorrhagic stage), whereas IgM is usually detectable seven days after symptom onset for a considerable time [12]. The sensitivity of these methods ranges from 75% to 100% for serological assays and 43% to 100% for molecular assays [13, 14]. However, CCHF lineages between geographical regions display the most significant degree of sequence diversity for any arbovirus, leading to a high risk of missed molecular detection [15]. In resource-poor settings, these diagnostic methods pose significant challenges. The absence of rapid, specific diagnostic tools further complicates timely identification and isolation, increasing the risk of hospital outbreaks and poor outcomes.

There are currently no approved vaccines or specific antiviral therapies, and supportive care remains the mainstay of management. Due to its high mortality, lack of effective countermeasures, and potential for widespread outbreaks, the World Health Organization has designated CCHF as a high-priority pathogen for research and development [16].

### Study rationale

This systematic review addresses the urgent need to evaluate simple clinical, laboratory, and epidemiological features associated with early diagnosis of Crimean-CCHF in resource-limited settings. We aim to synthesise data on patient characteristics that are significantly more or less prevalent in CCHF-positive individuals compared to controls, as reported in the medical literature.

By aggregating findings across multiple studies, this review seeks to improve the understanding of practical diagnostic indicators that can facilitate earlier recognition and management of CCHF, without proposing these as replacements for molecular or serological diagnostic methods. The findings are intended to inform clinical decision-making and public health strategies, particularly where laboratory infrastructure is constrained.

## Methods

### Protocol

This systematic review was conducted in accordance with the most recent Preferred Reporting Items for Systematic Reviews and Meta-Analyses (PRISMA) checklist [17]. This systematic review was officially registered with PROSPERO in June 2023 (ID: CRD42023434872). An official protocol was not otherwise produced for this review.

### Study eligibility

Studies were eligible if they reported data from patients suspected of CCHF, either based on clinical criteria or documented tick exposure. Only patients presenting to healthcare facilities in CCHF-endemic countries were included. An endemic country was defined as one with regularly reported (non-imported) cases [18]. Eligible studies recorded patient clinical signs/symptoms, laboratory findings, and/or epidemiological characteristics at admission. Measurement methods were not required to be specified for inclusion, but characteristics were required to either be measured routinely or to be feasible for low-resource settings. Data limited to highly specific biomarkers, viral load, or genetic polymorphisms were not considered.

The recorded characteristics of patients in eligible papers were separated between mutually exclusive groups of CCHF-positive and CCHF-negative patients in each study, with the latter acting as a control. The two groups ideally originated from the same source population, entering the study under the same eligibility criteria. Studies using healthy or asymptomatic controls were excluded. The outcome of interest was laboratory-confirmed CCHF, diagnosed by RT-PCR or IgM detection via ELISA. Diagnosis based solely on IgG was not considered sufficient. Further studies were excluded if they were not available in English, required payment for access, non-academic/peer reviewed, case reports, conducted in animals/cell lines or contained no primary data (mathematical modelling studies, reviews, and opinion articles). Studies without sufficient detail on the origin of their study population or data from comparison groups were also excluded. Observational and comparative studies providing primary data with full-text availability were included in this review. No date restrictions were applied.

### Study identification

The search was conducted in Embase, Global Health, MEDLINE, PubMed, and Cochrane Library, with the requirement for English language articles set. In addition, Google Scholar was also searched using a selection of keywords, namely ‘CCHF’, ‘early diagnosis’ and ‘clinical characteristics’.

The search began in April 2023 and is current up to June 10, 2023. The search remained the same across all databases used, except in the Ovid databases (Embase, MEDLINE and Global Health). The quotation marks here for “Crimean-Congo hemorrhagic fever” were removed to comply with Ovid search guidelines. The search consisted of three main components, consisting of multiple keywords and utilising truncations. Appropriate Boolean operators were used to link search terms as shown in Table 1. The search components covered the key aspects of CCHF, markers/indicators, and diagnosis respectively. MeSH terms were not used due to the possibility of excluding articles that have not yet been comprehensively indexed.

**Table 1.**
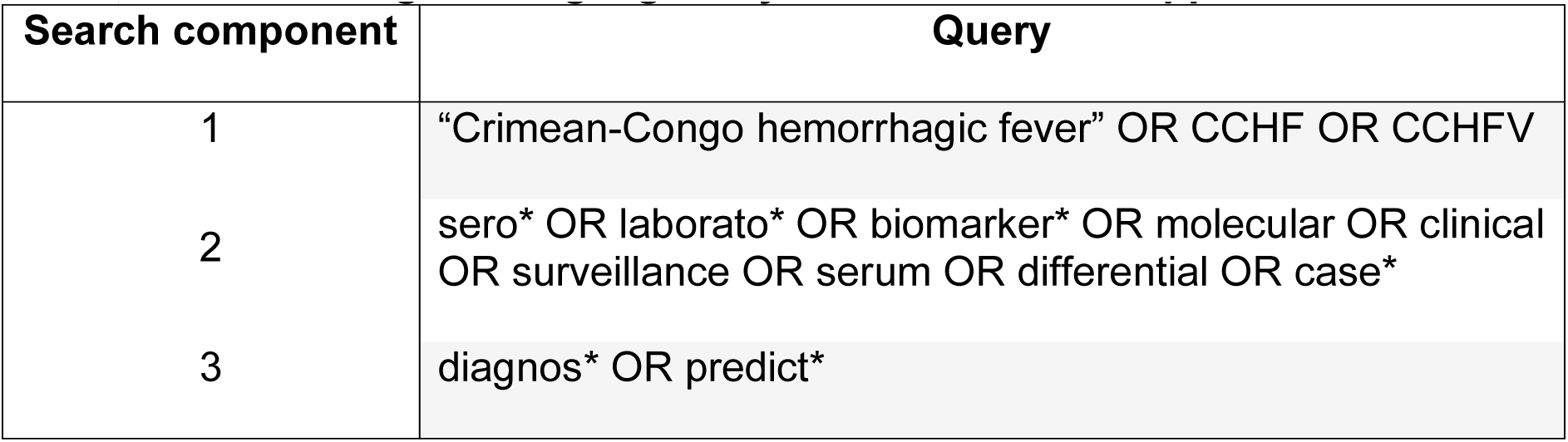
Search strategy – all three search components were combined using ‘AND’, a filter for English language only studies was then applied.

### Data extraction

The abstracts and titles of all articles were screened independently but in parallel by a first and second reviewer based on eligibility criteria, with studies deemed suitable then being retrieved. Full texts from this group were assessed for eligibility through two further parallel screenings. All citations were imported into Endnote 20 for screening, with duplicate studies also being removed. Fig 1 details the results of the literature search and study selection. Ethical approval was not required for this study as it takes the form of a literature review.

**Fig 1.**
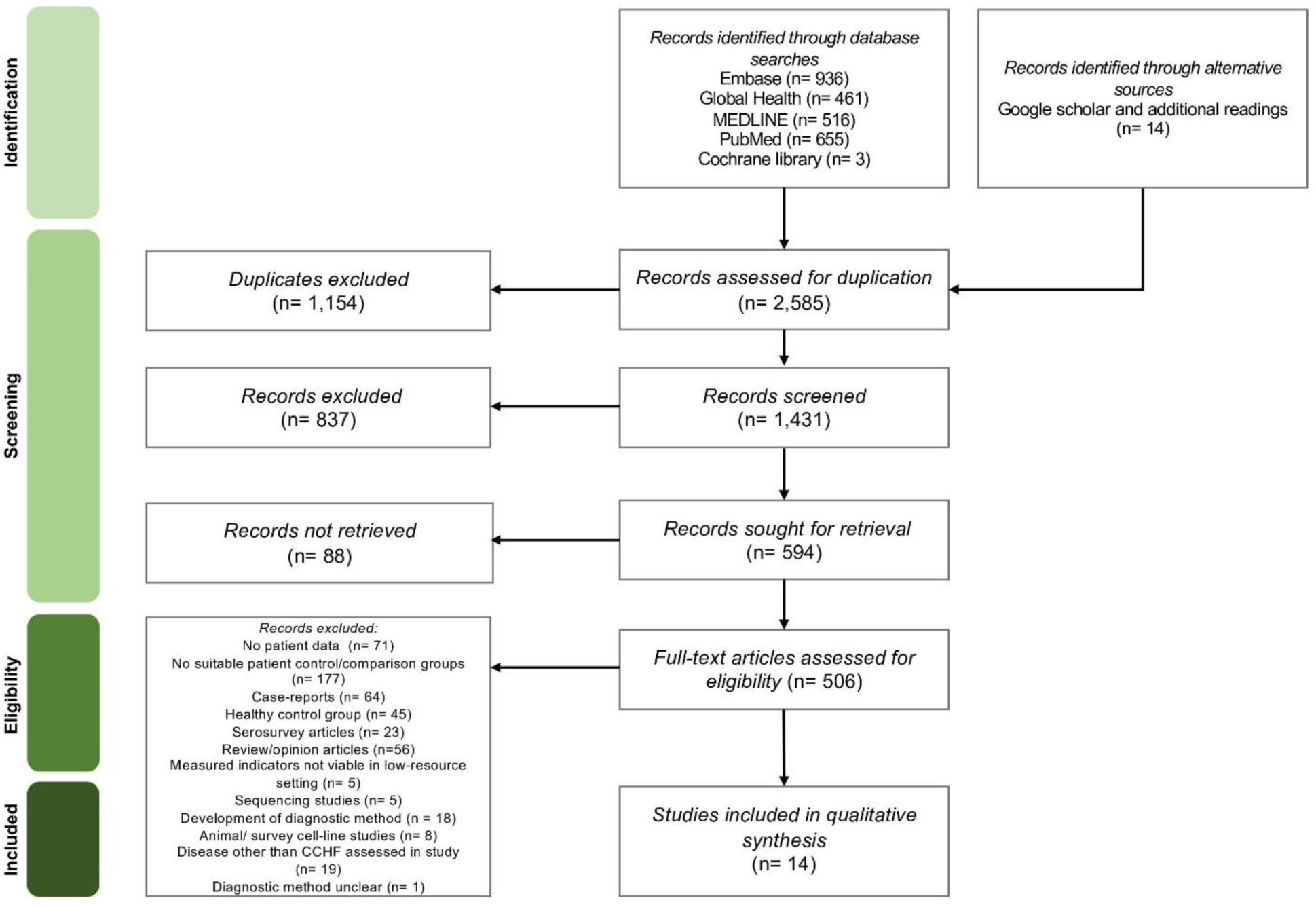
PRISMA flow diagram showing the process of study selection.

### Data synthesis

Gozalan et al. [19] and Karlberg et al. [20] did not provide descriptive statistical measures of association (p-values) for patient characteristics between the comparison groups. Accordingly, the author conducted Chi-squared tests for categorical variables and unpaired T-tests for continuous variables, using the extracted raw data on the frequency of each variable across CCHF-positive and CCHF-negative populations. A significance threshold of <0.05 was selected, in line with all other included studies. R statistical software was used for these tests and for the following calculations.

Odds ratios (ORs) and 95% Confidence Intervals (CIs) were calculated for all variables that were found at significantly different levels in CCHF-positive patients. These were conducted using the raw data on the listed frequencies of each recorded variable in each article. In instances where ORs could not be determined due to the presence of a zero-value (Kömürlüoğlu et al. [21] and Yeşilbağ et al. [22]), an adjustment of 0.5 was added to all groups to enable calculation. The necessary count data were not available in some articles. In these cases, means or medians were given instead as measures of central tendency. Odds ratios were not calculated in these cases. Other measures of effect size could also not be generated in these cases (see Discussion).

Forest plots were generated in Microsoft Excel for variables with more than three calculated ORs. A meta-analysis was considered, although due to qualitatively identified systematic issues regarding study design heterogeneity, pooled analysis or statistical measurement of heterogeneity was not deemed viable [23].

### Risk of bias assessment

Checklist tools from the Joanna Briggs Institute (JBI) were used to assess the risk of bias in each eligible study [24]. The case-control and cross-sectional tools were used, as both these designs were identified in the included studies. The case-control tool consists of ten questions (answered by yes, no, unclear, or N/A), followed by a decision on whether to include the study. The cross-sectional tool lists eight questions, most of which are similar to the case-control tool (Appendices 3 and 4). The determined bias level did not affect study inclusion, although the impact of these biases was later considered.

## Results

### Study selection

In total 2,571 records were identified through the five databases. A further 14 studies identified through Google scholar were also introduced at this point. After 1,154 duplicates were excluded from this number, the remaining 1,431 records had their abstracts and titles screened. After the initial screening, 594 articles were sought for retravel, with 88 found to be unavailable. The remaining 506 full-text articles were assessed for eligibility. A total of 14 studies were included in the full review and underwent data extraction. This process, and the reasons for study exclusion are shown below in Fig 1.

### Study characteristics

The characteristics of included studies are shown below in Table 1. Most studies (12 of 14) were conducted in Türkiye, with the remaining two being from Iran and India. These were all published between 2007-2021. The majority were retrospective case-control studies and just two were cross-sectional in design. A total of 2,025 participants were included in this review, with sample sizes ranging from 17 to 362. None of the study population cohorts used in each study were found to overlap with cohorts recruited in other included studies. Many studies included participants based on their own specific but similar criteria for a suspected CCHF patient (n=11) and the remaining were included based on recent history of tick bite (n=3). CCHF mortality ranged from 3.17-33.34%, with over half of studies listing a rate of under 10%.

### Critical appraisal and risk of bias assessment

Quality assessment summaries of the included studies using bias tools developed by the JBI are shown in Appendices 3 and 4 [24]. All 14 peer-reviewed articles had a clearly defined research objective; however, two studies did not aim to compare CCHF patient characteristics as their primary objective [31, 32].

### Clinical and methodological diversity

Several concerns were identified regarding the clinical diversity of study participants and their assessed patient characteristics. One key issue was the lack of consistency in case definitions. Four studies included patients suspected of CCHF based solely on tick-bite history (Table 2), whilst the remainder applied varying case definitions incorporating two or more non-specific symptoms (e.g., fever or nausea) in addition to a further laboratory or epidemiological signal. Three studies failed to clearly report these definitions [31, 33, 34]. Due to this heterogeneity, pooling odds ratios in a meta-analysis was deemed inappropriate [23]. However, the included studies fulfilled bias criteria enough to be accepted for qualitative analysis and findings should be interpreted in light of these inconsistencies.

**Table 2.** Characteristics of included studies.

| Study | Country | Study setting | Study design | Study period | Population | N of patient variables assessed | Most significant CCHF+ characteristics identified by authors | Sample size | CCHF diagnosis method | CCHF+ Mortality |
| --- | --- | --- | --- | --- | --- | --- | --- | --- | --- | --- |
| **Başol, Duygu and Ayan, 2013 [25]** | Türkiye | Emergency Department of the University Hospital of Tokat province | Case-Control | January - October 2012 | Patients admitted with tick bite | 4 | Fever, fatigue, leukopenia, thrombocytopenia, elevated AST/ALT | 251 (63 CCHF+, 188 CCHF-) | RT-PCR | 3.17% |
| **Mourya et al., 2017 [26]** | India | Various healthcare facilities within Gujarat and Rajasthan | Case-Control | January 2014 - September 2015 | Suspected CCHF patients (specific criteria) | 30 | Haemorrhagic manifestations including melaena, thrombocytopenia, elevated ALT | 69 (21 CCHF+, 48 CCHF-) | RT-PCR and IgM ELISA | 33.34% |
| **Erenler et al., 2015 [27]** | Türkiye | Emergency department of Hitit University Çorum Research and Education Hospital, Çorum province | Case-Control | January 2010 - December 2012 | Patients admitted with tick bite | 32 | Epistaxis, leukopenia, elevated CK/AST levels | 240 (97 CCHF+, 143 CCHF-) | RT-PCR | 21.65% |
| **Gozalan et al., 2007 [19]** | Türkiye | Various healthcare facilities within Tokat province | Case-Control | May 2002 - October 2003 | Suspected CCHF patients (specific criteria) | 18 | Epistaxis, leukopenia | 36 (29 CCHF+, 7 CCHF-) | RT-PCR or IgM ELISA | 3.45% |
| **Gozdas, 2019 [28]** | Türkiye | Inpatient clinic in Kastamonu State Hospital | Case-Control | 2014-2017 | Suspected CCHF patients (specific criteria) | 27 | Headache, nausea/vomiting, leukopenia, thrombocytopenia, elevated AST/ALT/LDH levels, tick bite, contact with animal material | 76 (31 CCHF+, 45 CCHF-) | RT-PCR or IgM ELISA | 9.68% |
| **Gürbüz et al., 2021** [29] | Türkiye | Various healthcare facilities within Gümüşhane province | Cross-Sectional | January 2011 - December 2019 | Suspected CCHF patients (specific criteria) | 35 | Headache, fever, diffuse bodily pain, nausea/vomiting, diarrhoea, tachycardia, elevated ALT/AST/CK/LDH levels, leukopenia, thrombocytopenia | 362 (242 CCHF+, 120 CCHF-) | RT-PCR or IgM ELISA | N/A |
| **Hekimoğlu and Demirci, 2014** [30] | Türkiye | Dr Münif İslamoğlu Hospital, Kastamonu province | Case-Control | 2013 | Suspected CCHF patients (specific criteria) | 27 | Tick bite and elevated CK levels | 41 (19 CCHF+, 22 CCHF-) | RT-PCR or IgM ELISA | 5.26% |
| **Karapinar et al., 2015** [31] | Türkiye | Cumhuriyet University Medical School, Sivas province | Case-Control | 2011-2012 | Suspected paediatric CCHF patients (specific criteria) | 22 | Thrombocytopenia | 54 (28 CCHF+, 26 CCHF-) | RT-PCR or IgM ELISA | - |
| **Karlberg et al., 2015** [20] | Iran | Boo-Ali Hospital, Sistan and Baluchestan province | Cross-Sectional | Not defined | Suspected CCHF patients (less specific criteria) | 8 | - | 17 (12 CHHF+, 5 CCHF-) | RT-PCR or IgM ELISA | - |
| **Kayadibi et al., 2019** [32] | Türkiye | Hitit University School of Medicine, Çorum province | Case-Control | 2014-2018 | Suspected CCHF patients (specific criteria) | 32 | - | 268 (149 CCHF+, 119 CCHF-) | RT-PCR | 3.36% |
| **Kılınc et al., 2016** [33] | Türkiye | Sabuncuoğlu Serafeddin Training and Research Hospital, Amasya province | Case-Control | 2011-2015 | Suspected CCHF patients (specific criteria) | 28 | Fever, headache, widespread body pain, fatigue, leukopenia, nausea, vomiting, elevated AST/ALT/LDH/CK levels, thrombocytopenia, absence of anaemia, tick bite, contact with animals, living in the countryside, animal husbandry | 281 (102 CCHF+, 179 CCHF-) | RT-PCR or IgM ELISA | 3.92% |
| **Kömürlüoğlu et al., 2017** [21] | Türkiye | Infectious Diseases Unit of Hacettepe University İhsan Doğramacı Children's Hospital, Ankara province | Case-Control | January 2014-December 2016 | Paediatric patients admitted with tick bite | 17 | - | 163 (6 CCHF+, 157 CCHF-) | RT-PCR or IgM ELISA | - |
| **Küçükceran et al., 2021** [34] | Türkiye | Emergency department of Erzurum regional training and research hospital | Case-Control | June - September 2012 | Suspected CCHF patients (specific criteria) | 24 | Leukopenia, thrombocytopenia, low fibrinogen levels, increased CK levels | 80 (40 CCHF+, 40 CCHF-) | RT-PCR or IgM ELISA | - |
| **Yeşilbağ et al., 2020** [22] | Türkiye | Yozgat state hospital, Yozgat province | Case-Control | 2011-2013 | Suspected CCHF patients (specific criteria) | 31 | Tick exposure, contact with animal material, leukopenia, anaemia | 87 (61 CCHF+, 26 CCHF-) | RT-PCR or IgM ELISA | 3.28% |

**RT-PCR** (-)
Reverse transcription-polymerase chain reaction Not available for retrieval

Study control groups were also inconsistently defined. One study [21] included non-symptomatic individuals and unbalanced comparison group sizes, leading to outlier odds ratios, which had to be excluded from the forest plots and were instead listed in the footnotes of Figs 2-4.

**Fig 2.**
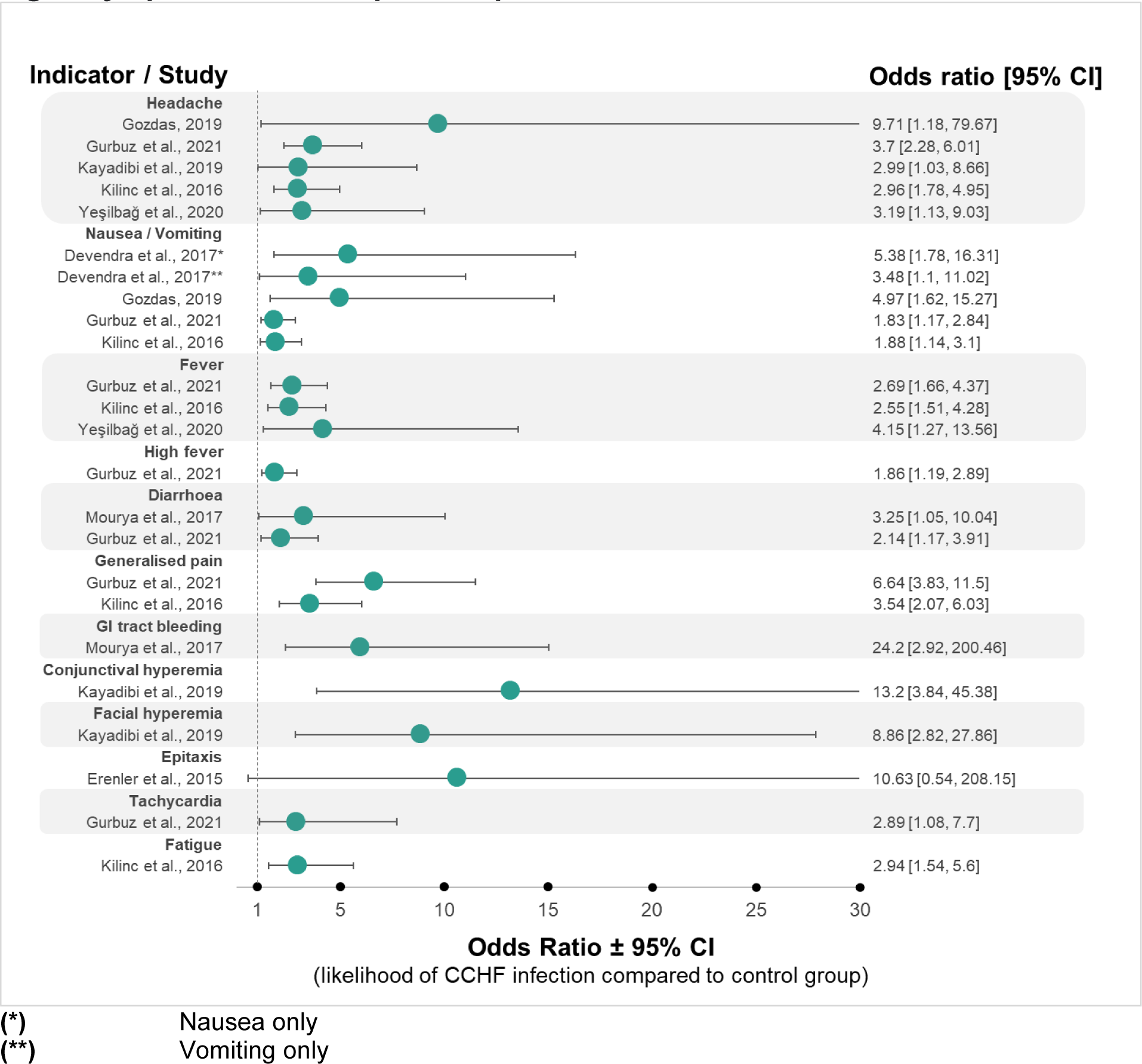
Forest plot of odds ratios for statistically significant clinical signs/symptoms of CCHF-positive patients identified. Findings from Kömürlüoğlu et al. [21] are listed here as the calculated ORs were deemed as outliers; Rash: 156 (95% CI: 12.36 – 1968.58), Fever: 228.65 (95% CI: 11.87 – 4405.12), Headache: 156 (95% CI: 12.36 – 1968.58), Vomiting: 125.83 (95% CI: 12.66 – 1251.17), Malaise: 256.67 (95% CI: 22.55-2921.75), Abdominal pain: 76.5 (95% CI: 10.71 – 546.58).

**Fig 3.**
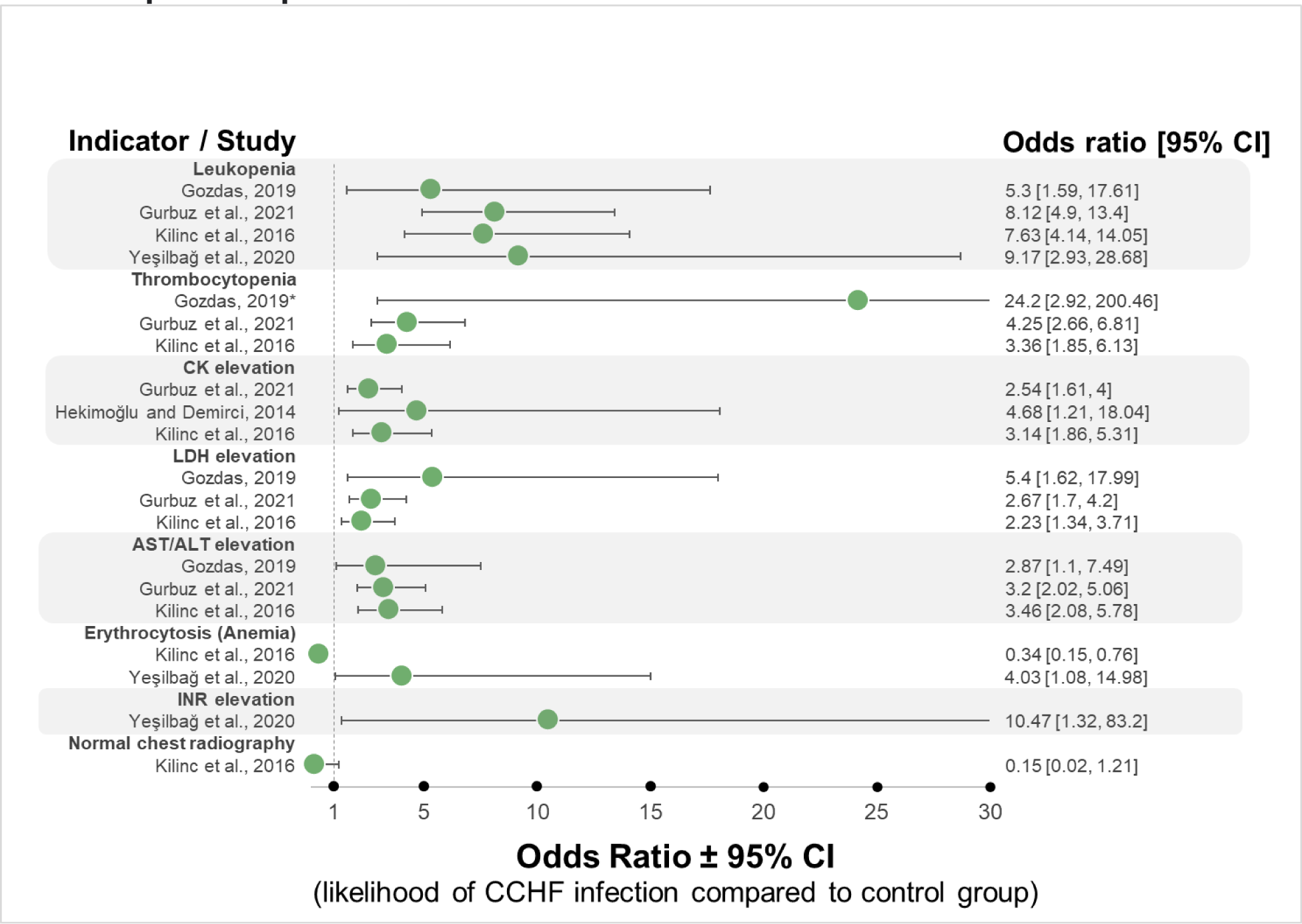
Forest plot of odds ratios for statistically significant laboratory findings of CCHF-positive patients.

**Fig 4.**
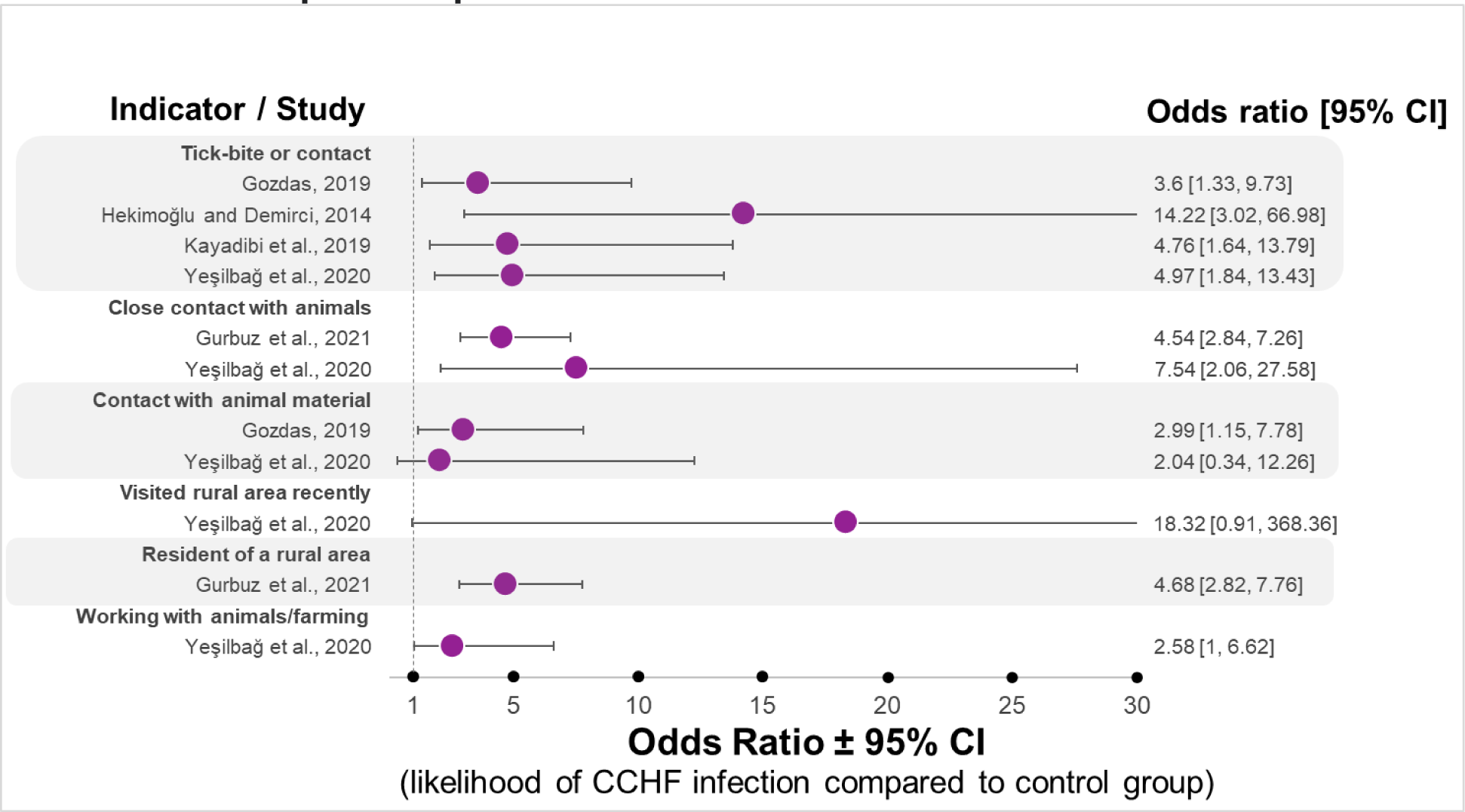
Forest plot of odds ratios for statistically significant epidemiological factors of CCHF-positive patients identified.

Other elements of study populations were poorly defined in some studies. For instance, Erenler et al. [27] screened a group of 15,429 patients admitted with tick-bite, but tested only 240 for CCHF without a clear rationale, suggesting selection bias. Similarly, Gozalan et al. [19] included only 36 out of 108 suspected cases with no clear explanation. Also, delays in serum sampling (up to 90 days post-symptom onset) in this study limit early-phase disease insight.

Methodological design diversity further compounds these issues. The retrospective design of several studies means they depend heavily on accurate medical record-keeping, introducing possible recall, reporting, and selection bias [19].

The effect of detection bias was reduced by requiring RT-PCR or IgM ELISA diagnosis, although variability in the length of the IgM detection window between patients could allow for false positives in patients still producing CCHF IgM for months after infection. Differences or overlap in symptom definitions, diagnostic thresholds, and unstandardised testing protocols may also have introduced measurement and misclassification bias.

Only three studies attempted to control for confounding via multivariate regression (Table 6). This means that whilst probable confounding of significant patient characteristics means causative links cannot be drawn, associations between characteristics and CCHF infection can still be observed, which still works to the advantage of the present aims. In addition, incomplete data limits odds ratio power in several studies [19, 26, 29].

Finally, publication bias likely influenced the included studies, as those with non-significant results may have been excluded from publication [35]. An extensive search spanning six databases was conducted as an attempt to compensate for the effect of these biases, although the overall findings may still lack generalisability or external validity.

### Summary of characteristics seen in CCHF-positive patients

Tables 3, 4, and 5 display CCHF-positive patient signs/symptoms, laboratory results and epidemiological factors found to be statistically significant from control groups in the analysed studies. Some similar or overlapping variables were combined to prevent repetition (Appendix 2). In addition, a laboratory finding denoted as ‘BK’ (p=0.019) was disregarded, as no explanation for its meaning could be found [21].

**Table 3.** Summary of significantly different clinical signs/symptoms between CCHF-positive and CCHF-negative patients.

| CLINICAL VARIABLES INCLUDED IN THE ANALYSIS | Number of studies included | Number of studies reporting a significant difference | % significant | Representative studies |
| --- | --- | --- | --- | --- |
| **CLINICAL SIGNS** |  |  |  |  |
| Rash | 7 | 1 | 14.29% | [21]*, [22], [28], [29], [30], [31], [33] |
| Epistaxis | 6 | 1 | 16.16% | [21], [22], [26], [27]*, [29], [33] |
| General bleeding | 6 | 0 | - | [19], [28], [30], [31], [32], [34] |
| Petechiae | 5 | 1 | 20% | [21]*, [22], [26], [29], [33] |
| Tachycardia | 5 | 1 | 20% | [22], [27], [29]*, [33], [34] |
| Hypotension | 5 | 0 | - | [22], [27], [29], [33], [34] |
| Gastrointestinal tract bleeding | 4 | 1 | 25% | [22], [26]*, [29], [33] |
| Gingival bleeding | 3 | 0 | - | [22], [29], [33] |
| Haematuria | 3 | 0 | - | [22], [29], [33] |
| Ecchymosis | 3 | 0 | - | [29], [33], [34] |
| Splenomegaly | 3 | 0 | - | [29], [31], [33] |
| High fever (>38 degrees Celsius) | 2 | 1 | 50% | [29]*, [31] |
| Conjunctival hyperaemia | 2 | 1 | 50% | [31], [32] |
| Hepatomegaly | 2 | 0 | - | [19], [31] |
| Facial hyperaemia | 1 | 1 | 100% | [32]* |
| Extended duration of fever | 1 | 0 | - | [31] |
| Tonsillopharyngitis | 1 | 0 | - | [31] |
| Jaundice | 1 | 0 | - | [26] |
| Anorexia | 1 | 0 | - | [26] |
| Nasal discharge | 1 | 0 | - | [21] |
| **CLINICAL SYMPTOMS** |  |  |  |  |
| Fever | 12 | 4 | 33.33% | [19], [21]*, [22]*, [26], [27], [28], [29]*, [30], [31], [32], [33]*, [34] |
| Headache | 11 | 6 | 55.55% | [19], [21]*, [22]*, [26], [28]*, [29]*, [30], [31], [32]*, [33]*, [34] |
| Nausea | 11 | 4 | 36.36% | [19], [22], [26]*, [27], [28]*, [29]*, [30], [31], [32], [33]*, [34] |
| Diarrhoea | 11 | 2 | 18.18% | [19], [22], [26]*, [27], [28], [29]*, [30], [31], [32], [33], [34] |
| Vomiting | 10 | 5 | 50% | [19], **[21]\***, [22], **[26]\***, **[28]\***, **[29]\***, [30], [32], **[33]\***, [34] |
| Abdominal pain | 9 | 1 | 11.11% | [19], **[21]\***, [26], [28], [29], [30], [31], [33], [34] |
| Altered level of consciousness | 8 | 0 | - | [19], [22], [26], [28], [29], [30], [33], [34] |
| Myalgia | 6 | 0 | - | [19], [26], [28], [30], [32], [34] |
| Fatigue | 4 | 1 | 25% | [22], [27], [28], **[33]\*** |
| Generalised pain | 3 | 2 | 66.67% | [22], **[29]\***, **[33]\*** |
| Arthralgia | 2 | 0 | - | [19], [26] |
| Weakness | 2 | 0 | - | [19], [30] |
| Malaise | 1 | 1 | 100% | **[21]\*** |
| Dizziness | 1 | 0 | - | [32] |
| Chills | 1 | 0 | - | [26] |
| Breathlessness | 1 | 0 | - | [26] |
| Sore throat | 1 | 0 | - | [31] |
| Vertigo | 1 | 0 | - | [32] |

\* Statistically significant difference in indicator between CCHF-positive and CCHF-negative patients (p <0.05)

**Table 4.**
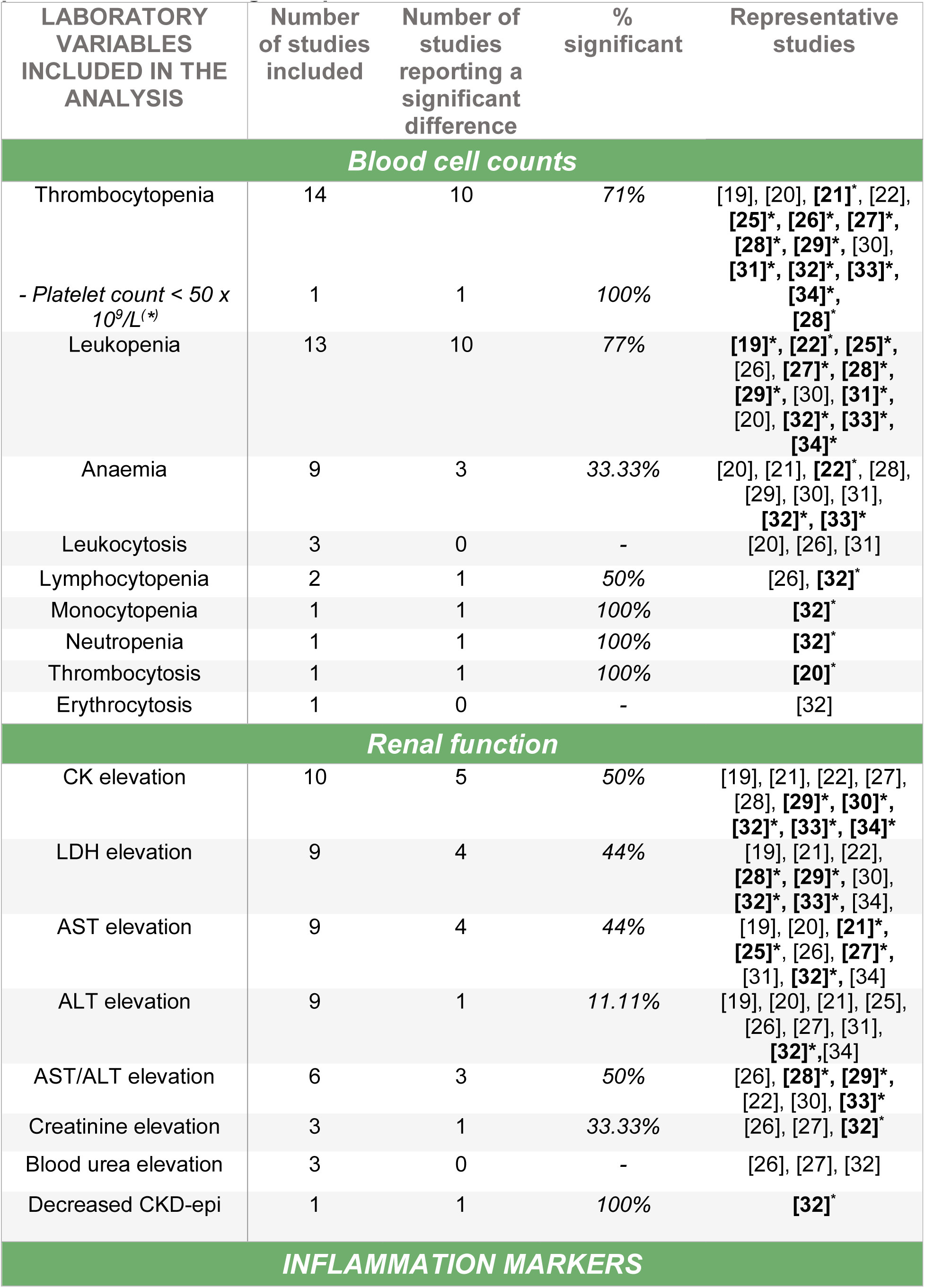
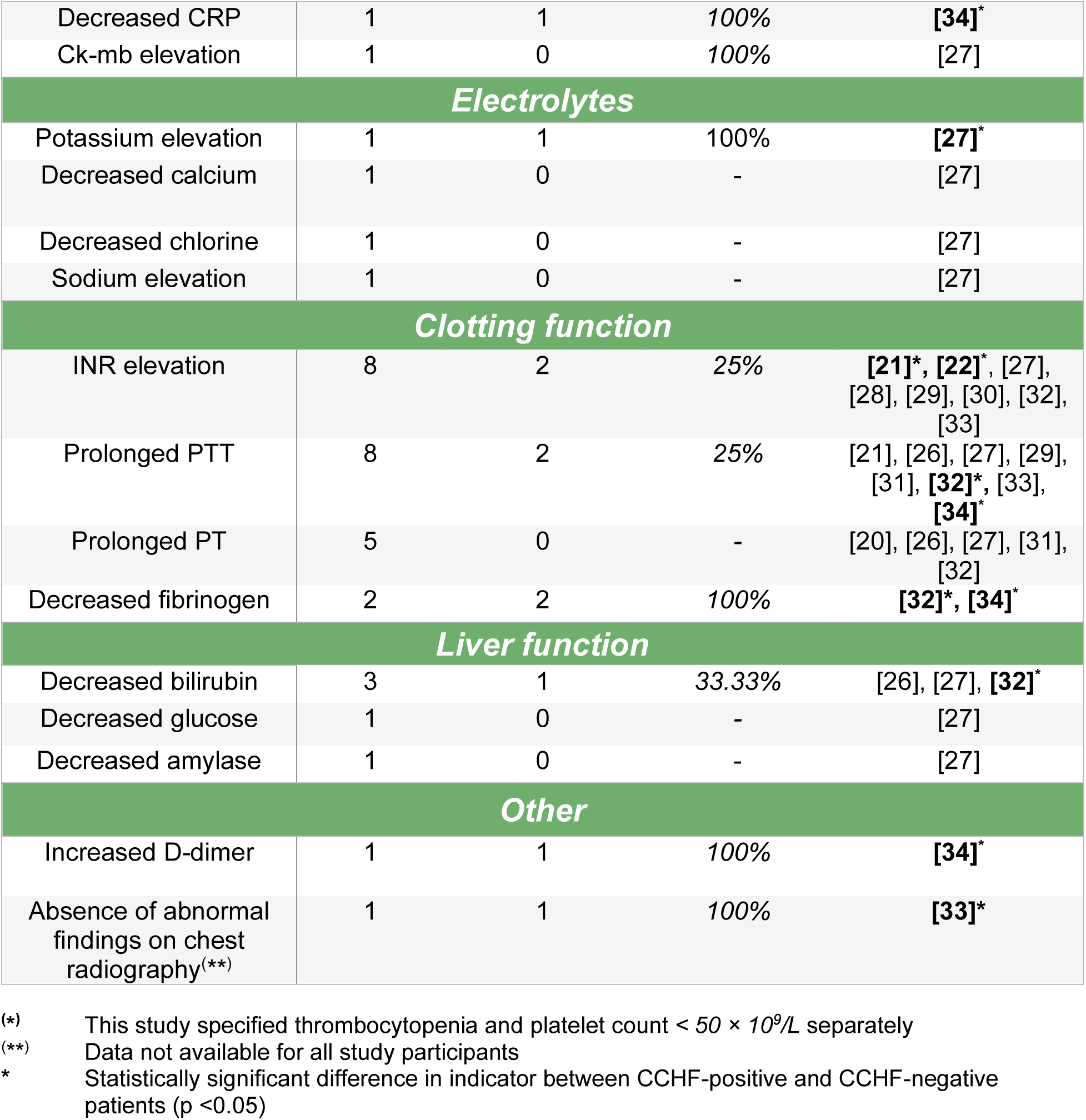
Summary of significantly different laboratory findings between CCHF-positive and CCHF-negative patients.

The most frequently identified significant clinical symptoms were fever (n=4), headache (n=6), and nausea/vomiting (n=5). No individual clinical signs were reported as significantly more/less common in CCHF-positive patients more than twice. The most common laboratory findings indicative of a CCHF-positive diagnosis were elevated LDH (n=4), elevated CK (n=5), transaminitis (n=7), leukopenia (n=10), and thrombocytopenia (n=10). In terms of epidemiological factors, tick bite/contact (n=4) was most commonly identified in CCHF-positive patients.

Odds ratios for each significantly different patient characteristic are displayed in Figs 2-4. All listed significant patient characteristics were found in higher amounts in CCHF-positive patients, apart from ‘Normal findings on chest radiographic’ in Fig 3, where the CCHF patients were less likely to display this. Those frequently identified significant CCHF patient characteristics all scored consistently high ORs. Leukopenia was especially indicative of CCHF diagnosis.

Studies also reported several other variables for which near-significant differences (p ≤ 0.1) between CCHF-positive and CCHF-negative groups were found. These were: Mourya et al. [26] (abdominal pain p=0.06, anorexia p=0.086), Gozalan et al. [19] (myalgia p=0.086), Karapinar et al. [31] (extended duration of fever p=0.053, AST elevation p=0.103), Kayadibi et al. [32] (erythrocytosis p=0.07, diarrhoea p=0.098, decreased INR p=0.052), Kılınç et al. [33] (tachycardia p=0.051, decreased INR p=0.07), Küçükceran et al. [34] (myalgia p=0.105, raised AST p=0.105, decreased creatinine p=0.067) and Yeşilbağ et al. [22] (generalised pain p=0.07).

Three studies also conducted multivariate logistic regression analysis to account for confounding variables [22, 26, 31]. A summary of the variables that were found to be independently predictive of CCHF diagnosis are shown in Table 6. Many characteristics remained non-significant between CCHF-positive and CCHF-negative patients across the included studies. The most prominent of these similarities, were altered consciousness (n=8), general bleeding (n=6), myalgia (n=6), prolonged PT (n=5), hypotension (n=5), gingival bleeding (n=3), haematuria (n=3), splenomegaly (n=3), ecchymosis (n=3), blood urea elevation (n=3), leukocytosis (n=3) and contact with a CCHF case (n=3). Those with very few significant differences were epistaxis (n=1 of 6), rash (n=1 of 7), tachycardia (n=1 of 5), petechiae (n=1 of 5), abdominal pain (n=1 of 9), diarrhoea (n=2 of 11), ALT elevation (n=1 of 9), and living in a rural area (n=1 of 5).

**Table 5.** Summary of significantly different epidemiological factors between CCHF-positive and CCHF-negative patients.

| EPIDEMIOLOGICAL VARIABLES INCLUDED IN THE ANALYSIS | Number of studies included | Number of studies reporting a significant difference | % significant | Representative studies |
| --- | --- | --- | --- | --- |
| **Events** |  |  |  |  |
| Tick bite or contact | 4 | 4 | 100% | [22]*, [28]*, [30]*, [32]* |
| Close contact with animals | 4 | 2 | 50% | [22]*, [28], [29]*, [30] |
| Contact with animal material | 3 | 2 | 66.66% | [22]*, [28]*, [30] |
| Visited rural area recently | 3 | 1 | 33.33% | [22]*, [28], [30] |
| Contact with CCHF cases | 3 | 0 | - | [22], [28], [30] |
| **TICK REMOVAL** |  |  |  |  |
| Self-removal | 2 | 0 | - | [22], [27] |
| At hospital | 1 | 0 | - | [27] |
| Suspected bite | 1 | 0 | - | [27] |
| **PLACE OF RESIDENCE** |  |  |  |  |
| Rural area | 5 | 1 | 20% | [22], [28], [29]*, [30], [32] |
| Town centre | 2 | 0 | - | [27], [29] |
| Village | 1 | 0 | - | [27] |
| City | 1 | 0 | - | [27] |
| **OCCUPATION** |  |  |  |  |
| Farming/animal husbandry | 3 | 1 | 33.33% | [22]*, [29], [32] |
| Undeclared | 1 | 0 | - | [29] |
| Student or government worker | 1 | 0 | - | [29] |
| Child (<16 yr) | 1 | 0 | - | [29] |
| Healthcare worker | 1 | 0 | - | [29] |

\* Statistically significant difference in indicator between CCHF-positive and CCHF-negative patients (p <0.05)

**Table 6.** Summary of variables found to be independently predictive of CCHF diagnosis in multivariate regression.

|  | Odds ratio | 95% CI | P-value |
| --- | --- | --- | --- |
| ***Mourya et al . [26]*** |  |  |  |
| PLT count (x10<sup>9</sup>/L) | 0.05 | 0.006 – 0.412 | - |
| Melaena | 14.6 | 1.3 – 163.5 | - |
| Nausea | 9.1 | 1.3 – 64.2 | - |
| ***Kayadibi et al. [32]*** |  |  |  |
| Conjunctival hyperaemia | - | - | - |
| Direct bilirubin (mg/dL) | <0.001 | <0.001 – 0.001 | <0.001 |
| Lymphocytes (10<sup>3</sup>/uL) | 0.005 | 0.001 – 0.046 | <0.001 |
| Fibrinogen (mg/dL) | 0.979 | 0.970 – 0.988 | <0.001 |
| AST (U/L) | 1.042 | 1.021 – 1.064 | <0.001 |
| CKD-EPI (mL/min/1.73m<sup>2</sup>) | 0.931 | 0.898 – 0.966 | <0.001 |
| Haematocrit (%) | 1.398 | 1.173 – 1.665 | <0.001 |
| Neutrophils (10<sup>3</sup>/uL) | 0.588 | 0.439 – 0.787 | <0.001 |
| Creatine kinase (U/L) | 0.999 | 0.997 – 1.000 | 0.033 |
| ***Yeşilbağ et al. [22]*** |  |  |  |
| Tick exposure | 9.03 | 1.96 – 41.47 | 0.005 |
| Contact with animal material | 14.92 | 2.23 – 99.94 | 0.005 |
| Leukopenia | 13.65 | 2.55 – 72.91 | 0.02 |
| Anaemia | 8.41 | 1.06 – 66.42 | 0.04 |

These analyses were conducted by and retrieved from their respective articles.
(-) Not provided by the article

## Discussion

### Key findings

This review identified numerous potential predictors of CCHF, with leukopenia and thrombocytopenia the most prominent. Other frequent predictors included transaminitis, headache, nausea/vomiting, elevated CK, fever, LDH elevation, and tick bites. Multivariate analyses from several studies supported strong independent associations between CCHF diagnosis and thrombocytopenia, leukopenia, nausea, and tick/animal exposure, while links with elevated CK/AST showed possible confounding.

Paediatric cases of CCHF are thought to present differently from adult cases, typically following a milder course [36], although the present study could not confirm this. The ages of study populations for most studies were poorly defined and often only provided a mean age, making it unclear if paediatric cases were considered or in what quantity. This is with the exception of three articles [20, 30, 34], which all specified that children were excluded. Conversely, Karapinar et al. [31] and Kömürlüoğlu et al. [21] only included paediatric cases. Interestingly, Karlberg et al. [20] was the only study to note a significantly higher prevalence of thrombocytosis in CCHF-positive patients, whilst Hekimoğlu and Demirci [30] may have shown a comparatively higher OR for tick bite/contact (Fig 4). Kömürlüoğlu et al., [21] showed comparatively much higher ORs across all potential predictors of CCHF, however this is much more likely due to the inequitable study population of CCHF-positive and CCHF-negative patients. It has been suggested that perceived differences in the clinical presentation of young children may be since they are less likely to self-report their symptoms [37]. In addition, undifferentiated fevers are much more common in childhood, meaning milder disease is more likely to go unnoticed. The present study is unable to make a judgment on whether children do indeed present differently, due to the small and heterogeneous comparisons.

The inclusion criteria used for the included study populations were highly heterogeneous between studies, which may affect comparison. In addition, population demographics such as age were poorly defined. It is unclear whether this led to any observable differences in patient characteristics.

### Implications for research

Previous studies have identified similar findings in CCHF patients to those presented in this review [12, 38]. A case study in Uganda [39] found that the most commonly reported CCHF symptoms were fever (93.8%), haemorrhage (81.3%), headache (78.1%), fatigue (68.8%), vomiting (68.8%), and myalgia (65.6%). Other varied abnormalities were also observed, including thrombocytopenia, leukopenia, and anaemia. Most participants also reported tick bites or exposure to livestock as their potential source of infection. These findings are encouraging, as they indicate the findings of the present study may be generalisable outside of the Middle East. Additional studies support the presence of these findings as strongly suggestive of CCHF diagnosis [40, 41, 42].

Predictive symptoms of Ebola virus disease (EVD) have been examined in the literature [43], although this study focused on rapid recognition during large outbreaks rather than diagnosis of sporadic cases in resource-limited settings. Pooled odds ratios were calculated, identifying a range of symptom-based predictors (rather than any laboratory indicators). Several early, non-specific symptoms were strongly associated with EVD, including diarrhoea (pOR 2.99), fatigue (pOR 2.77), vomiting (pOR 2.69), fever (pOR 1.97), muscle pain (pOR 1.65), and cough (pOR 1.63). By contrast, the present review found stronger predictors for CCHF, particularly laboratory-based markers such as leukopenia and thrombocytopenia.

High-risk populations in low-income countries would benefit from a Rapid Diagnostic Test (RDT) for CCHF, and an antigen-based lateral flow test showing high sensitivity (92.9%) and specificity (96.2%) is in development [44]. However, similar projects have faced slow progress toward regulatory approval and distribution [45].

While RDTs/POC testing remain the ultimate goal, interim improvements can be made by enhancing recognition of CCHF-specific symptoms, supporting diagnosis in resource-limited settings. Even if successfully developed, RDT distribution to remote communities could take years, highlighting the need for complementary approaches such as standardised diagnostic algorithms. Importantly, the findings of this review are not offered as an alternative to novel diagnostics, but as a bridge until access to RDTs improves.

### Limitations

Several limitations should be considered when interpreting the findings of this review. Most included studies (14 of 16) were conducted in Türkiye, limiting generalisability outside of this region. This is especially true regarding distant regions, where CCHF presentations may differ due to variations in viral lineage, although the extent of this effect is unclear [46]. Additionally, many studies spanned long recruitment periods, during which viral mutations may have occurred, reducing comparability across time (see Table 2) [29, 32, 33]. Many studies were also conducted in secondary or tertiary hospitals, introducing referral filter bias, and reducing the representativeness of primary care settings.

The diagnostic utility of the present findings is further complicated by the absence of reliable exclusion in study populations for common co-infections. Infections such as malaria and dengue are important differential diagnoses for CCHF and share many clinical similarities, most importantly thrombocytopenia and leukopenia [47, 48]. Early clinical presentations are also very similar, including headache, myalgia, nausea, and vomiting. This may confound the present study’s findings in malaria- and dengue-endemic regions. While testing for these infections is more widely available, tick exposure can help differentiate CCHF; however, misclassification remains a concern.

Many continuous variables lacked qualitative assessment. The use of a mean or standardised mean difference to compare the predictive abilities of continuous variables was considered but deemed not feasible due to inconsistent laboratory protocols and missing individual-level data, making the requisite calculation of the population standard deviation impossible. This limits deeper interpretation of continuous predictors.

The analysis in this review also relied heavily on p-values, which can misrepresent associations in smaller studies [49, 50]. While odds ratios with 95% confidence intervals were also included, large effect sizes seen in Figs 2-4 may not reflect true associations. Several key findings are based on a small subset of studies, increasing the risk of bias. Furthermore, four studies had fewer than 50 participants, reducing the overall statistical power of this review.

Diagnostic access bias may also affect this review, as all studies confirmed CCHF through PCR or ELISA, tools often unavailable in rural LICs. This may limit the applicability of findings in resource-constrained settings. Similarly, the emphasis on laboratory-based findings as predictors may reduce clinical utility where such tools/equipment are lacking. The exclusion of non-English language studies may also have introduced selection bias to this review.

Clinical and laboratory findings likely varied by stage of infection in the study populations. Whilst this adds heterogeneity, it reflects real-world conditions where patients will likely present to healthcare centres at different points in illness progression. Despite these limitations, this review provides a meaningful synthesis of the available evidence. It is important, though, that its conclusions not be generalised beyond the context of CCHF cases presenting to healthcare facilities.

## Conclusion

The findings presented in this review suggest that easily identifiable predictors of CCHF-positive patients are potentially viable to inform patient triage for clinicians. Leukopenia and thrombocytopenia were especially indicative of CCHF diagnosis. Characteristics found to be very similar between CCHF-positive and CCHF-negative patients may also aid in differential diagnosis. These findings could serve as interim guidance until sensitive diagnostic technologies are more widely available in remote and low-resource areas.

Empirical evidence comparing similarly presenting CCHF-positive and CCHF-negative patients across multiple studies was previously lacking prior to this review. Whilst it is hoped the present findings may be applied to healthcare settings and provide diagnostic utility, they should be interpreted with caution due to unclear generalisability and study heterogeneity. Increased detailed reporting and surveillance of CCHF incidence will aid future clinical assessments. It is hoped that this study will inform clinicians and policymakers, aiding in the differentiation of CCHF-positive and similarly presenting patients visiting healthcare facilities.

## Data Availability

Data underlying the results in the study are within the manuscript and Supporting Information Files.

## Supporting information

**S1 File. Appendix**. Appendix 1. Fields searched in databases, Appendix 2. Combinations of similar variables in present study, Appendix 3. Risk of bias assessment of Case-Control studies JBI tool, Appendix 4. Risk of bias assessment of Cross-sectional studies using JBI tool, Appendix 5. Suspected CCHF case definitions (inclusion criteria) as described in included studies.

**S2 File. Data from included studies.** Dataset of patient clinical signs/symptoms, laboratory results, and epidemiological factors as recorded in the included studies.

**S3 File. Completed PRISMA checklist.** PRIMSA 2020 checklist with relevant page numbers and manuscript excerpts added.

## Notes

### Competing Interest Statement

The authors have declared no competing interest.

### Funding Statement

The author(s) received no specific funding for this work.

### Author Declarations

No ethnical approval was required as this study was a systematic review.

